# Integrated multiomic scores for cardiovascular disease prediction across multiancestral populations

**DOI:** 10.64898/2026.10.23.26366372

**Authors:** Pascal B. Meyre, Michael Chong, Marie Pigeyre, Shihong Mao, Michael Kamal, Shrikant Bangdiwala, Roya Kelishadi, Annika Rosengren, Fernando Lanas, Afzalhussein Yusufali, Rita Yusuf, Jephat Chifamba, Patricio Lopez-Jaramillo, Marc Evans M. Abat, Alvaro Avezum, Karen Yeates, Maria Luz Diaz, Hassan Mir, Bernard Keavney, Sonia S. Anand, Koon Teo, Sumathy Rangarajan, Philip Joseph, Hertzel Gerstein, Salim Yusuf, Guillaume Paré

**Author notes:** Corresponding author: Guillaume Paré, MD, MSc, McMaster University, Population Health Research Institute, David Braley Cardiac, Vascular and Stroke Institute, 237 Barton St. East, Room C3103, Hamilton, Ontario L8L 2X2, Canada,.

## Abstract

**Background:** Genomic, epigenomic, and proteomic data capture complementary aspects of cardiovascular disease biology, but the value of integrating these data for predicting multiple cardiovascular disease outcomes across ancestry groups remains uncertain.

**Methods:** We derived disease-specific proteomic scores using 999 plasma proteins in 50,673 UK Biobank participants and validated them in 9,358 participants from the Prospective Urban and Rural Epidemiological (PURE) study. In PURE, we integrated these proteomic scores with 2,279 polygenic scores and 479 DNA methylation scores to develop 4 multiomic scores (MEGASCOREs) for myocardial infarction (MI), atrial fibrillation (AF), heart failure (HF), and cardiovascular mortality. Associations with incident cardiovascular disease, discrimination, calibration, and performance across ancestry groups were evaluated in PURE and 2 additional cohorts.

**Results:** In pooled analyses, each standard deviation increase in the corresponding MEGASCORE was associated with MI (hazard ratio [HR], 2.13; 95% CI, 1.86-2.43), AF (HR, 1.94; 95% CI, 1.68-2.24), HF (HR, 3.33; 95% CI, 2.75-4.05), and cardiovascular mortality (HR, 3.28; 95% CI, 2.33-4.61). MEGASCOREs improved discrimination compared with clinical models, with strongest improvements for HF and cardiovascular mortality. MEGASCORE_CV-Death_ had a higher area under the receiver operating characteristic curve (AUC) than SCORE2 for cardiovascular mortality (0.83 [95% CI, 0.82-0.85] vs. 0.78 [95% CI, 0.77-0.80]). Its discrimination was also higher than that of SCORE2 across all 4 ancestry groups, with better calibration across non-European ancestry groups. MEGASCOREs remained associated with cardiovascular outcomes among participants without conventional cardiovascular risk factors.

**Conclusions:** Integrated multiomic MEGASCOREs were associated with multiple cardiovascular outcomes and provided prognostic information beyond conventional clinical variables in PURE.

**Clinical Perspective:** *What is new?:*

- We developed 4 cardiovascular disease-specific multiomic scores integrating proteomic signatures, polygenic scores, and DNA methylation scores to characterize risk of MI, AF, HF, and cardiovascular mortality.
- The MEGASCOREs were associated with incident cardiovascular disease outcomes and provided prognostic information beyond conventional clinical risk factors.
- MEGASCOREs showed consistent risk stratification across ancestry and remained associated with risk among participants without conventional cardiovascular risk factors.

*What are the clinical implications?:*

- Integrated multiomic scores may complement conventional cardiovascular risk assessment by capturing molecular information not fully represented by clinical variables.
- Molecular risk profiling may help identify individuals with elevated cardiovascular risk despite having no conventional risk factors.
- Assessment of clinical utility, evaluation of implementation and cost-effectiveness are needed before MEGASCOREs can inform preventive treatment decisions.

## Introduction

Recent advances in genomic, epigenomic, and proteomic profiling have expanded opportunities to characterize the molecular processes underlying cardiovascular disease.^1–3^ These data capture complementary dimensions of risk. Polygenic and methylation-based scores quantify inherited susceptibility and cumulative environmental exposures,^4–6^ whereas circulating proteomic profiles provide a dynamic snapshot of active pathophysiological processes.^7^ Integrating these molecular layers may improve cardiovascular risk stratification beyond conventional clinical variables and help characterize the biological heterogeneity underlying different cardiovascular diseases.

Previous studies have demonstrated that multiomic approaches can improve risk stratification beyond single-modality scores.^8–13^ However, the extent to which genomic, epigenomic, and proteomic scores provide complementary information across multiple cardiovascular diseases remains incompletely understood. In particular, the transportability of integrated multiomic scores across populations with diverse ancestry and clinical characteristics requires further evaluation. Establishing whether these scores improve discrimination and risk stratification relative to established clinical prediction models is an important step toward assessing their potential clinical relevance.

In this study, we developed 4 cardiovascular disease-specific multiomic scores, called MEGASCOREs, for myocardial infarction (MI), atrial fibrillation (AF), heart failure (HF), and cardiovascular mortality. We derived proteomic scores in the UK Biobank and integrated these with polygenic and DNA methylation scores in the multinational PURE study. We evaluated associations with incident cardiovascular disease outcomes, predictive performance relative to clinical models, and risk stratification among participants without conventional cardiovascular risk factors. We further assessed the performance of the MEGASCORE across ancestry groups and evaluated its transportability in 2 additional cohorts.

## Methods

### Cohorts

The UK Biobank (UKB) is a prospective cohort of >500,000 individuals aged 40-69 year recruited between 2006 and 2010.^14^ The UK Biobank-Pharma Proteomics Project (UKB-PPP) is a nested subset of participants with plasma proteomic profiling performed using Olink platforms.^15^ Of 54,219 participants with proteomic data, 50,673 remained after quality control exclusions. A total of 999 proteins overlapped with those measured in the validation cohort (PURE study) and were retained for analyses. Outcome-specific proteomic scores were derived for all-cause mortality, cardiovascular mortality, MI, HF, AF, chronic kidney disease, type 2 diabetes, and stroke. Outcomes were ascertained through linkage to hospital records, primary care data, death registries, and self-reports, and harmonized using three-digit ICD codes. Detailed outcome definitions are provided in the **Supplementary methods**. All participants provided informed consent, and the UKB received ethics approval from its ethics oversight committee.

The Prospective Urban Rural Epidemiology (PURE) study is a multinational, multiethnic prospective cohort with approx. 225,000 participants aged 35 to 70 years from more than 27 high, middle and low-income countries.^16^ Detailed description of the study design is provided in the **Supplementary methods**. A subset participated in a biobanking initiative for genomic, epigenomic and proteomic profiling. Blood samples from 14 countries were processed under standardized protocols and analyzed at the Population Health Research Institute (Hamilton, Canada). We implemented a case-cohort design, selecting a random subcohort from 55,246 eligible participants, supplemented by all additional participants who developed outcomes of interest (**Supplementary Figure 1**). After exclusion of samples failing genomic, epigenomic, or proteomic quality control or with missing/extreme values, the final analytic cohort included 9,358 individuals. Outcome events were prospectively collected using the definitions, similar to that used in the UKB. The study was approved by research ethics committees at each study centre and at Hamilton Health Sciences (Hamilton, ON, Canada), and all study participants provided written informed consent.

The Global Congestive Heart Failure (G-CHF) is a prospective, observational registry of patients with clinically diagnosed HF. The cohort enrolled 23,341 HF patients between 2016 and 2020 from 257 outpatient and hospital inpatient centers across 40 countries on 5 continents.^17,18^ Adults aged ≥18 years were eligible if they had a clinical diagnosis of HF based on the judgment of the local investigator. A subset of patients was invited to participate in a molecular substudy. The substudy included 2,431 participants with available baseline plasma for proteomic assays and whole-blood DNA for genetic and epigenetic profiling. Study outcomes included cardiovascular and non-cardiovascular mortality, as well as cardiovascular disease outcomes including MI and AF. Patients were followed every 6 months up to 6 years (annual clinic visits and telephone calls at 6 and 18 months), with assessment of vital status and hospitalizations at each contact. The study was approved by the local ethics committees, and all participants provided written informed consent.

The Outcomes Reduction with an Initial Glargine Intervention (ORIGIN) trial was a randomized study of 12,537 individuals with dysglycemia (prediabetes or diabetes) who were assigned to insulin glargine or standard care and, independently, to omega-3 fatty acids or placebo in a 2×2 factorial design.^19,20^ Participants were followed for a median of 6.2 years, during which the incidence and date of several cardiovascular-related outcomes were recorded. A subset of patients participated in the molecular substudy. A total of 323 participants had available baseline plasma for proteomic assays and whole-blood DNA for genetic and epigenetic profiling. Several health outcomes were analyzed according to their prior adjudication status during the trial. These included cardiovascular mortality, fatal or nonfatal MI, and HF. The trial was approved by the ethics review board at each participating center, and all participants provided written informed consent.

### Development and validation of proteomic scores

Details of proteomic profiling in all cohorts are provided in the **Supplementary methods**. In UKB and PURE, proteins with <20% missing values were imputed using *k*-nearest neighbors imputation.^21^ Expression values (NPX) were rank-based inverse normal-transformed to ensure approximate normality and standardized to a mean of 0 and standard deviation (s.d.) of 1 prior to statistical modeling. In UKB, eight outcome-specific proteomic scores were derived in the for all-cause mortality, cardiovascular mortality, MI, stroke, HF, AF, type 2 diabetes, and chronic kidney disease. Proteins associated with each outcome were selected using least absolute shrinkage and selection operator (LASSO) regression.^22^ Model performance was assessed by the area under the receiver operating characteristic curves (AUC). The resulting UKB-derived protein coefficients were applied to the corresponding protein measurements in PURE to calculate the proteomic scores. The exact outcome definition, prediction horizon, and method used to derive predicted probabilities should be specified in the **Supplementary methods**.

### Construction and external validation of MEGASCOREs

MEGASCOREs were developed in PURE using proteomic scores derived in the UKB, together with publicly available polygenic risk scores and DNA methylation risk scores. Details of genomic and epigenomic profiling in PURE, G-CHF, and ORIGIN are provided in the **Supplementary methods**. Polygenic risk scores were obtained from the PGS Catalog using summary statistics from genome-wide association studies (GWAS) available through May 2025.^23^ Methylation risk scores were generated from epigenome-wide association studies (EWAS) using summary statistics for epigenome-wide significant CpG sites listed in the EWAS Catalog over the same period.^24^ A total of 2,279 polygenic and 479 methylation risk scores were generated for each PURE participant. Using these scores together with the 8 UKB-derived proteomic scores, four cardiovascular disease-specific MEGASCOREs were constructed for MI (MEGASCORE_MI_), HF (MEGASCORE_HF_), AF (MEGASCORE_AF_) and cardiovascular mortality (MEGASCORE_CV-Death_) by combining the corresponding component scores with LASSO regression. To minimize overfitting, we used a 5-fold cross-validation framework in which four folds were used for model training and the remaining fold for validation in each iteration (**Supplementary Figure 2**). The resulting MEGASCOREs were standardized to a mean of 0 and an s.d. of 1.

We evaluated the transportability of the four MEGASCOREs for MI, HF, AF and cardiovascular mortality in G-CHF and ORIGIN. Proteomic scores were calculated by applying the fixed coefficients from the UKB-derived proteomic models to the corresponding protein measurements. Polygenic and methylation risk scores were calculated using the same methods as described above, where the required data were available. For each MEGASCORE, the previously established component-specific weights and coefficients were applied, and the component scores were combined using the fixed coefficients from the PURE-derived MEGASCORE model. The final MEGASCOREs were standardized to a mean of 0 and a s.d. of 1.

### Statistical analysis

Pairwise correlations between MEGASCOREs were assessed using Pearson correlation coefficients. Because of the small sample size, selected analyses were not performed in ORIGIN. Event rates of MI, AF, HF and cardiovascular mortality were calculated across MEGASCORE deciles over the available follow-up. Associations between MEGASCOREs and cardiovascular disease outcomes were evaluated using multivariable Cox proportional hazards models. Hazard ratios (HRs) and 95% confidence intervals (CIs) were estimated per 1-s.d. increase in the score and across score quartiles. In PURE, the case-cohort sampling design was accounted for using the Self-Prentice method. Models were adjusted for age, sex, ancestry (African + Middle Eastern [Arab, Persian], Asian, European, or Latin American), body mass index (BMI), smoking (current/former smoker or never smoker), height, weight, hypertension, diabetes, and hypercholesterolemia. The proportional hazards assumption was assessed using Schoenfeld residuals, with no violations observed. Cohort-specific HRs per s.d. increase in MEGASCORE were pooled using inverse-variance-weighted random-effects meta-analysis. AF was not ascertained in ORIGIN, and incident HF was not assessed in G-CHF. Kaplan-Meier curves were generated to visualize event-free survival across MEGASCORE quartiles.

Discrimination was evaluated using AUCs for 4 model specifications: (1) age and sex; (2) cardiac disease-specific clinical model; (3) MEGASCORE alone; and (4) combined clinical model plus MEGASCORE. For MI, the clinical model included age, sex, BMI, smoking, hypertension, diabetes, and hypercholesterolemia. For AF, HF, and cardiovascular mortality, validated clinical scores were used, including CHARGE-AF, Framingham Heart Failure score (FHS HF score), and SCORE2.^25–27^ Model discrimination was compared using DeLong tests. Calibration was assessed by comparing predicted and observed risks across MEGASCORE deciles and estimating calibration slopes and intercepts. For cardiovascular mortality, the performance of MEGASCORE_CV-Death_ was compared with that of SCORE2 across the 4 ancestry groups.

Prespecified subgroup analyses examined event rates across MEGASCORE deciles among participants without conventional cardiovascular risk factors at baseline. Participants were classified as having no conventional risk factors if they had no obesity (BMI ≥30 kg/m²), smoking, hypertension, diabetes or hypercholesterolemia. Models in this subgroup were adjusted for age, sex, ancestry, and BMI. Further analyses characterized MEGASCOREs in relation to clinical, socioeconomic, and lifestyle factors and investigated their biological correlates. Associations between MEGASCOREs and lifestyle, socioeconomic, and cardiovascular risk factors were assessed using multivariable linear regression adjusted for age, sex, ancestry, and all risk factors. Effect estimates were reported as standardized regression coefficients and corresponding *t*-statistics. Standardized NT-proBNP levels were compared across quartiles of MEGASCORE_AF_ and MEGASCORE_HF_, with trends assessed across quartiles. MEGASCOREs were evaluated according to adherence to a healthy lifestyle. Healthy lifestyle factors included non-smoking, BMI <30 kg/m², high physical activity based on International Physical Activity Questionnaire,^28^ and a healthy diet, defined as being in the top tertile of the Alternative Healthy Eating Index.^29^ Participants were classified as having a favorable lifestyle (≥3 healthy factors), as having a favorable lifestyle (≥3 healthy factors), an intermediate lifestyle (2 factors), or an unfavorable lifestyle (≤1 factor). Differences in MEGASCOREs across lifestyle categories were assessed using multivariable linear regression adjusted for age, sex and ancestry. Associations between MEGASCORE tertiles (low, intermediate, high) and cardiovascular disease outcomes were evaluated using multivariable Cox models adjusted for cardiovascular risk factors and the relevant cardiovascular disease-specific clinical scores, with the intermediate tertile as the reference group. Adjusted Kaplan-Meier estimates were used to generate cumulative incidence curves. Gene Ontology enrichment analysis of proteins included in each proteomic score was performed using over-representation analysis in *clusterProfiler* (v4.0). Unsupervised *k*-means clustering was used to classify participants according to their four MEGASCOREs. Incidence rates of cardiovascular disease outcomes were estimated per 100 person-years across clusters. Cumulative incidence curves and Cox models were used to evaluate associations between clusters and a composite cardiovascular disease outcome comprising MI, AF, HF, or cardiovascular mortality. All analyses were performed using R version 4.4. All statistical tests were two-sided, with P<0.05 considered statistically significant.

## Results

Using 999 plasma proteins measured in both UKB and PURE, we derived and validated proteomic scores for 8 diseases and mortality (**Figure 1a, Supplementary Tables 1-8, Supplementary Figure 3**). We then integrated 2,279 polygenic risk scores, 479 methylation risk scores, and 8 proteomic scores in PURE to construct four cardiovascular disease-specific multiomic MEGASCOREs for MI, HF, AF, and cardiovascular mortality. The scores showed stability across cross-validation folds (**Supplementary Figures 4 and 5**).

**Figure 1.**
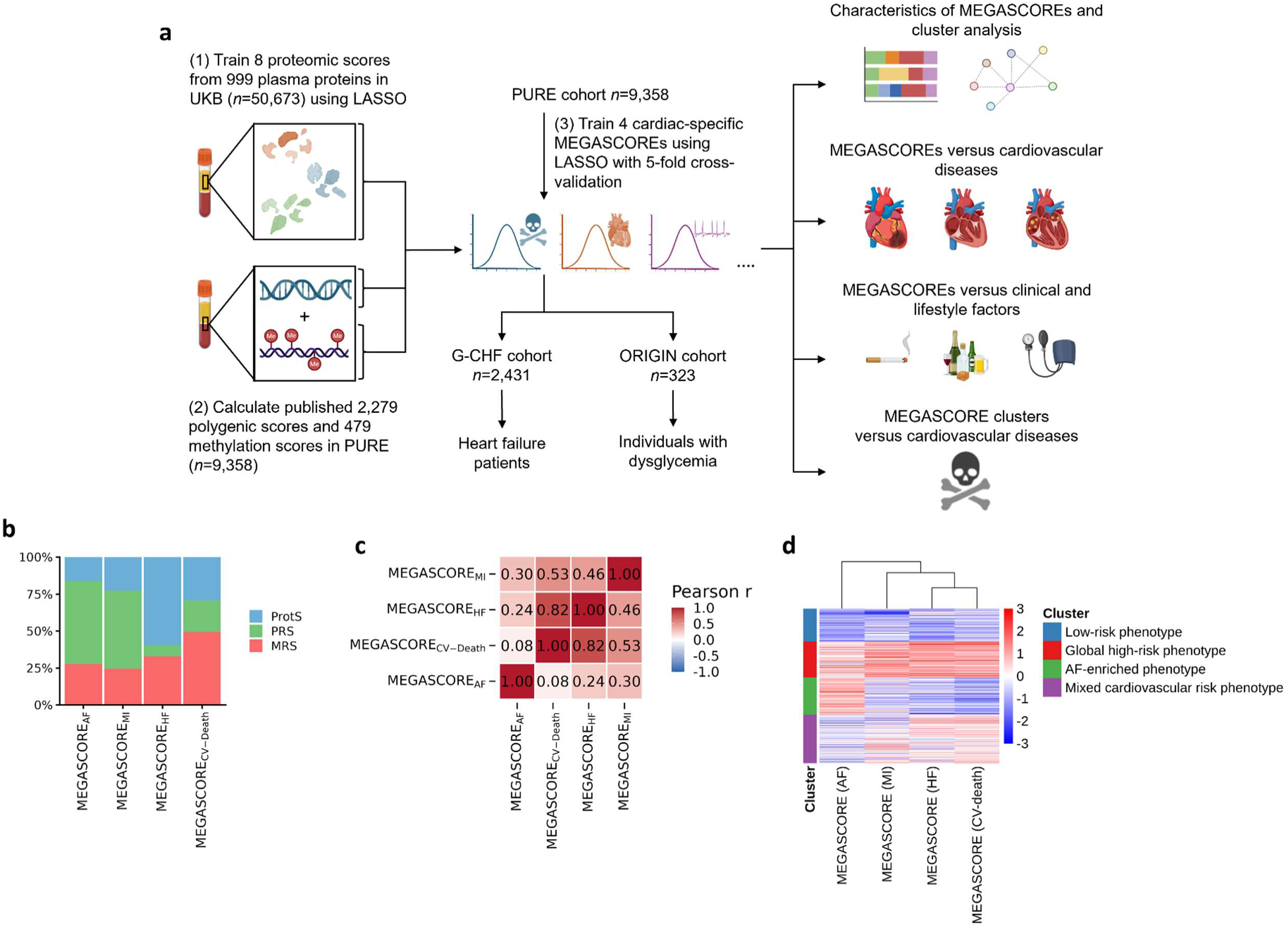
Derivation and composition of the MEGASCOREs. **a**, Study design to derive disease-specific proteomic scores from plasma protein data in the UKB and the construction of integrated, multi-omic, cardiac disease-specific MEGASCOREs in PURE. Eight outcome and cardiometabolic disease-specific proteomic scores for all-cause mortality, cardiovascular mortality, MI, stroke, heart failure, atrial fibrillation, diabetes, and chronic kidney disease from 999 plasma proteins were trained in 50,673 UKB participants using LASSO and externally validated in 9,358 PURE participants. Proteomic scores were then combined with 2,279 calculated polygenic scores and 479 methylation scores, both calculated from genome- and epigenome wide association studies summary statistics. MEGASCOREs were calculated using LASSO regression with nested five-fold cross-validation and then z-standardized to mean of 0, and 1 s.d. MEGASCOREs were validated in two independent cohorts; G-CHF includes 2,431 patients with diagnosed heart failure and ORIGIN includes 323 patients with dysglycemia. Downstream analyses assessed associations of MEGASCOREs with incident cardiac diseases, clinical and lifestyle factors, pathway enrichment, and mortality. **b**, Proportional contribution of proteomic, polygenic, and methylation component scores within each cardiac-specific MEGASCORE. **c**, Correlation matrix of the four cardiac disease-specific MEGASCOREs, illustrating shared and distinct molecular architectures. **d**, Unsupervised clustering of PURE individuals based on z-values of the four MEGASCOREs. Graph was created with BioRender.com. Abbreviations: AF=atrial fibrillation, MI=myocardial infarction, HF=heart failure, CV=cardiovascular, UKB=UK Biobank, PURE=Prospective Urban and Rural Epidemiological, LASSO= Least Absolute Shrinkage and Selection Operator, G-CHF=Global Congestive Heart Failure, ORIGIN=Outcomes Reduction with an Initial Glargine Intervention.

### MEGASCOREs for cardiovascular disease and individual compositions

The MEGASCORE_AF_ incorporated 12 omic scores, the MEGASCORE_MI_ included 17, MEGASCORE_HF_ included 19, and the MEGASCORE_CV-Death_ 28 scores. Polygenic risk scores constituted the largest proportion of component scores included in the MEGASCORE_AF_ (56%) and MEGASCORE_MI_ (53%), whereas proteomic scores accounted for the largest proportion included in the MEGASCORE_HF_ (60%) (**Figure 1b**). Methylation scores constituted the largest proportion of component scores in MEGASCORE_CV-Death_ (49%). Pairwise correlations between MEGASCOREs were modest overall, with the strongest correlation observed between MEGASCORE_HF_ and MEGASCORE_CV-Death_ (Pearson *r*=0.80; **Figure 1c**). Unsupervised clustering of MEGASCOREs identified 4 distinct molecular profiles in PURE: A low-risk cluster with uniformly low scores (n=1,996; 21%), a global high-risk cluster characterized by elevated scores across all MEGASCORE domains (n=2,160; 23%), and two disease-enriched clusters characterized by predominant AF-related signatures (AF-enriched; n=2,267; 24%) or mixed cardiovascular risk (mixed cardiovascular-enriched; n=2,994; 32%) (**Figure 1d**).

### Prediction of MEGASCOREs for cardiovascular diseases across cohorts and among individuals without conventional risk factors

Baseline characteristics of participants in PURE, G-CHF and ORIGIN are shown in the **Supplementary Table 9**. MEGASCOREs were associated with their corresponding disease outcomes after adjustment for major cardiovascular risk factors (**Figure 2a**). The pooled HR/s.d. in z-standardized MEGASCORE_CV-Death_ was 3.28 (95% CI, 2.33-4.61) for cardiovascular mortality. Similarly, MEGASCORE_MI_ (pooled HR/s.d. = 2.13; 95% CI, 1.86-2.43), MEGASCORE_AF_ (pooled HR/s.d. = 1.94; 95% CI, 1.68-2.24), and MEGASCORE_HF_ (pooled HR/s.d. = 3.33; 95% CI, 2.75-4.05) were each significantly associated with their respective outcomes. Kaplan-Meier curves further demonstrated progressively lower survival across MEGASCORE_CV-Death_ quartiles in G-CHF, ORIGIN, and PURE (**Figure 2b**). In multivariable-adjusted Cox regression, all four MEGASCOREs showed graded associations with their respective outcomes across quartiles in G-CHF, ORIGIN, and PURE (**Figure 2c**). Area under the ROC curve (AUC) analyses showed that individual MEGASCOREs improved discrimination of AF, MI, HF, and cardiovascular mortality compared with clinical models (**Figure 2d and Supplementary Figure 6**). For cardiovascular mortality, the MEGASCORE_CV-Death_ achieved an AUC of 0.83 (95% CI, 0.82-0.85), compared with 0.78 (95% CI, 0.77-0.80) for SCORE2 in PURE (P_DeLong_ =3.21×10^−27^). A similar comparison in G-CHF showed AUCs of 0.70 (95% CI, 0.68-0.73) for the MEGASCORE_CV-Death_ and 0.57 (95% CI, 0.54-0.60) for SCORE2 (P_DeLong_ =2.24×10^−30^). The MEGASCOREs showed acceptable calibration (**Supplementary Figure 7**).

**Figure 2.**
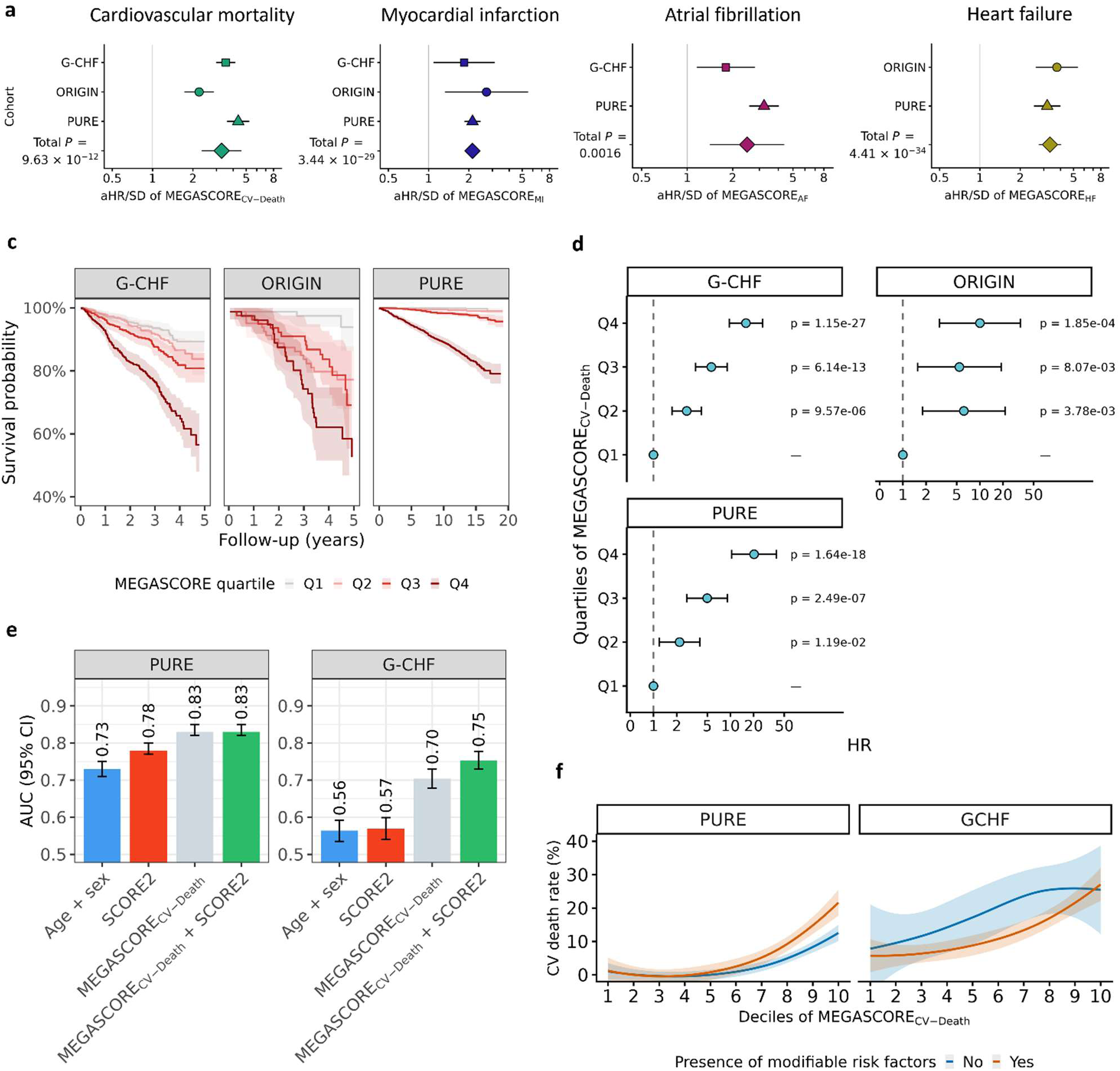
Prediction of future cardiovascular disease by MEGASCOREs. **a**, Random-effects meta-analysis on the associations between cardiac disease specific MEGASCOREs per s.d. and future cardiovascular diseases and mortality across cohorts from multivariable adjusted Cox regression analyses. Multivariable models are adjusted for age, sex, body-mass index (BMI), smoking, height, weight, hypertension, diabetes, and hypercholesterolemia. **b**, Kaplan-Meier curves with 95% confidence intervals for cardiovascular mortality across quartiles of MEGASCORE_CV-Death_ in the G-CHF, ORIGIN, and PURE cohorts. **c**, Association between MEGASCORE_CV-Death_ quartiles and future cardiovascular mortality from Cox regression analyses. Estimates are presented as hazard ratios with 95% confidence intervals from multivariable-adjusted models. **d**, Predictive ability of SCORE2, MEGASCORE_CV-Death_ and their combination in predicting cardiovascular mortality using area under the ROC (AUC). **e**, Event rates of cardiovascular mortality according to MEGASCORE deciles in individuals with at least one and without conventional risk factors in PURE and G-CHF. Estimates are percentages and are unadjusted. Abbreviations: SCORE2=Systematic Coronary Risk Evaluation 2.

MEGASCOREs remained associated with incident cardiovascular diseases among those without cardiovascular risk factors in PURE (n=3,394) and G-CHF (n=361), with event rates increasing progressively from the lowest to the highest MEGASCORE decile (**Figure 2e**). In both cohorts, the MEGASCORE_CV-Death_ improved prediction of cardiovascular mortality in individuals without risk factors (**Supplementary Figure 8**). The addition of clinical variables to the MEGASCORE minimally improved AUC values.

### Cross-ancestry comparison of MEGASCORE_CV-Death_ and SCORE2 for prediction of cardiovascular mortality

We compared the performance of MEGASCORE_CV-Death_ with SCORE2 for predicting cardiovascular mortality across ancestry groups in PURE. MEGASCORE_CV-Death_ demonstrated consistent risk stratification across all ancestry groups, with progressively higher observed cardiovascular mortality rates across increasing score deciles (**Figure 3a**). In contrast, SCORE2 showed greater variability in cardiovascular mortality rates across ancestries, particularly among European participants. Discrimination of cardiovascular mortality was superior for MEGASCORE_CV-Death_ compared with SCORE2 in all ancestry groups (**Figure 3b**). The AUCs of MEGASCORE_CV-Death_ were 0.84 in African and Middle Eastern, 0.78 in Asian, 0.86 in European, and 0.84 in Latin Americans, compared with corresponding AUCs of 0.80, 0.75, 0.78, and 0.76 for SCORE2. Calibration analyses demonstrated good agreement between predicted and observed cardiovascular mortality across ancestries for MEGASCORE_CV-Death_, whereas SCORE2 showed miscalibration in non-European groups, including Asian, African and Middle Eastern, and Latin Americans (**Figure 3c**).

**Figure 3.**
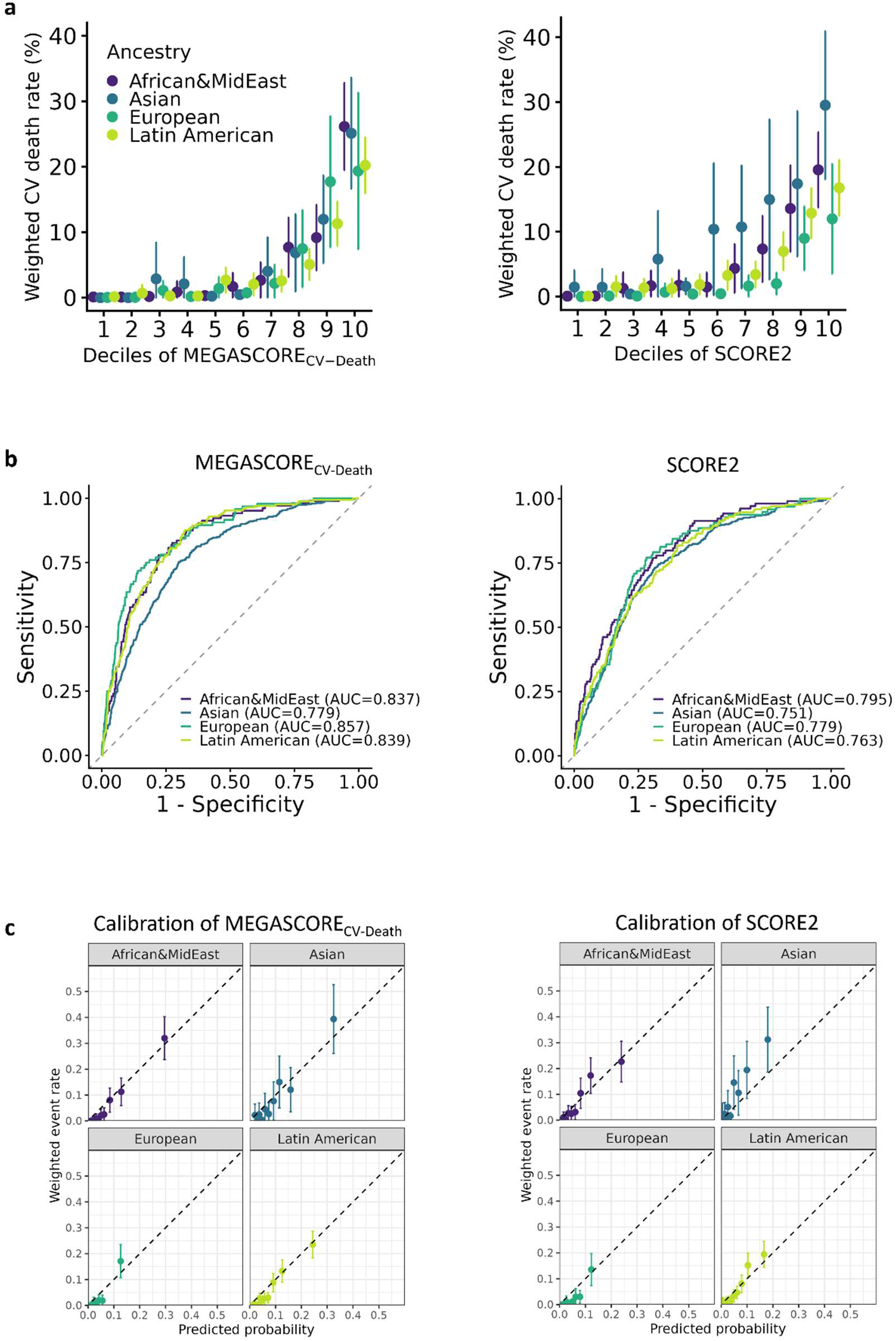
Cross-ancestral comparison of MEGASCORE_CV-Death_ and SCORE2 for prediction of cardiovascular mortality in PURE. **a**, Cardiovascular mortality rates across deciles of MEGASCORE_CV-Death_ and SCORE2, stratified by ancestry. Estimates are percentages and are unadjusted. **b**, Discrimination of MEGASCORE_CV-Death_ and SCORE2 for predicting cardiovascular mortality shown as area under the ROC (AUC) across ancestry groups. **c**, Calibration of MEGASCORE_CV-Death_ and SCORE2 for cardiovascular mortality across ancestry groups. Predicted versus observed risk are shown across score deciles.

### Association of MEGASCOREs with clinical and lifestyle factors

We next tested the sensitivity of MEGASCOREs to lifestyle, socioeconomic and cardiovascular risk factors in PURE. We found 14 positive and 11 negative significant associations (P<0.05). Smoking, alcohol, and high stress levels were associated with higher MEGASCORE values, whereas higher physical activity and education were associated with lower MEGASCOREs (**Figure 4a**). Hypertension and diabetes were associated with higher MEGASCORE_HF_ and MEGASCORE_CV-Death_, and hypercholesterolemia showed the strongest association with MEGASCORE_AF_ (**Figure 4b**). Individuals in the highest quartile of MEGASCORE_AF_ or MEGASCORE_HF_ showed higher standardized NT-proBNP values compared to those in the lowest MEGASCORE quartiles (**Figure 4c**). Compared with an unfavorable lifestyle, favorable and intermediate lifestyle profiles were associated with lower MEGASCORE values, particularly for MEGASCORE_MI_ and MEGASCORE_CV-Death_, whereas differences in MEGASCORE_AF_ were small (**Figure 4d**).

**Figure 4.**
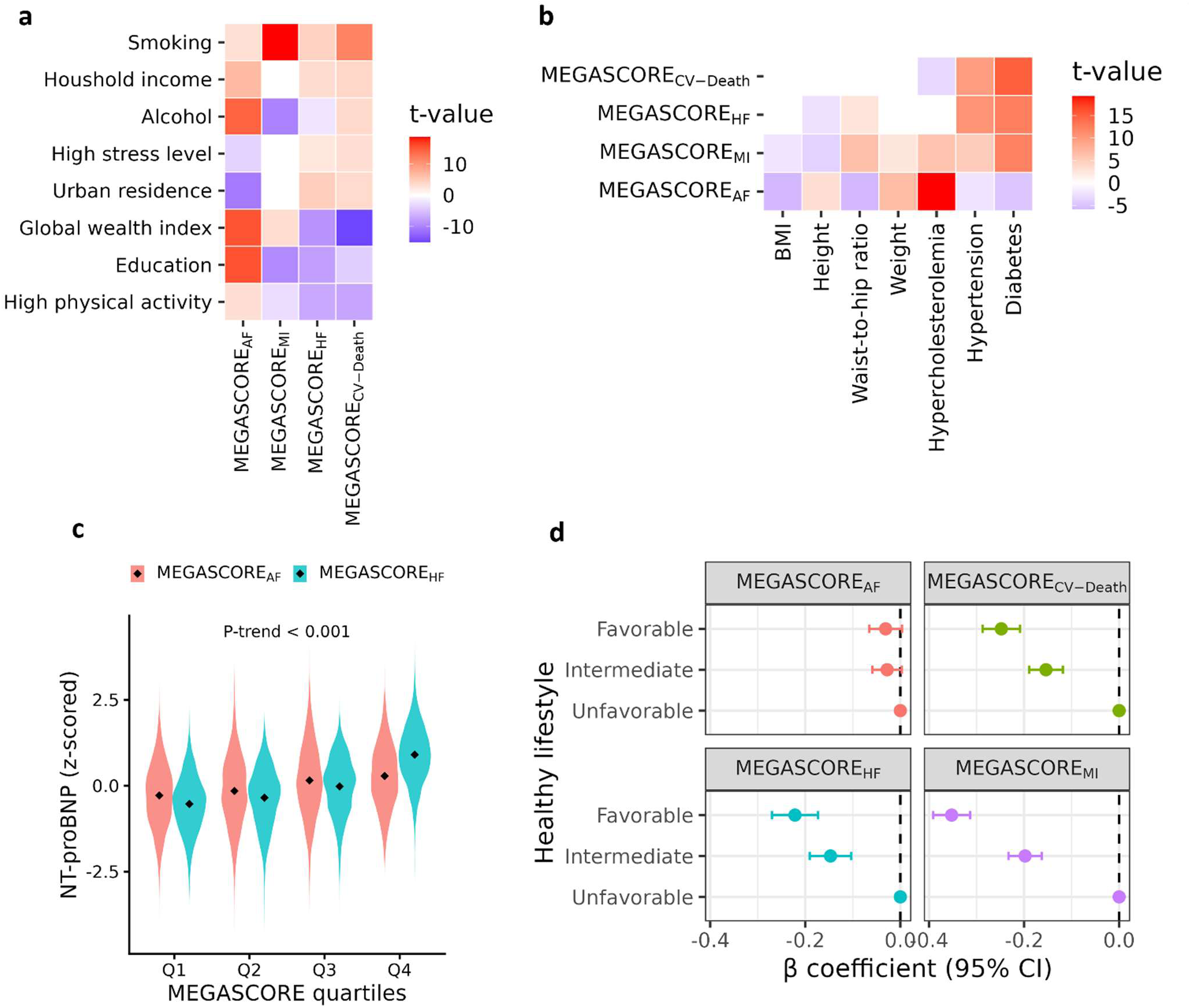
Association of MEGASCOREs with clinical and lifestyle factors. **a,b** Associations between cardiac disease-specific MEGASCOREs and socioeconomic and lifestyle factors. Heatmaps show t-values from linear regression models relating each MEGASCORE to socioeconomic, lifestyle and cardiometabolic risk factors. **c**, Violin plots of z-standardized NT-proBNP values across quintiles of the MEGASCORE_AF_ and MEGASCORE_HF_. Violin plots display median and interquartile range, with whiskers indicating 1.5×IQR. **d**, Linear regression for the association between MEGASCOREs and healthy lifestyle status (favorable, intermediate and unfavorable). Models are adjusted for age, sex and ancestry.

**Figure 5.**
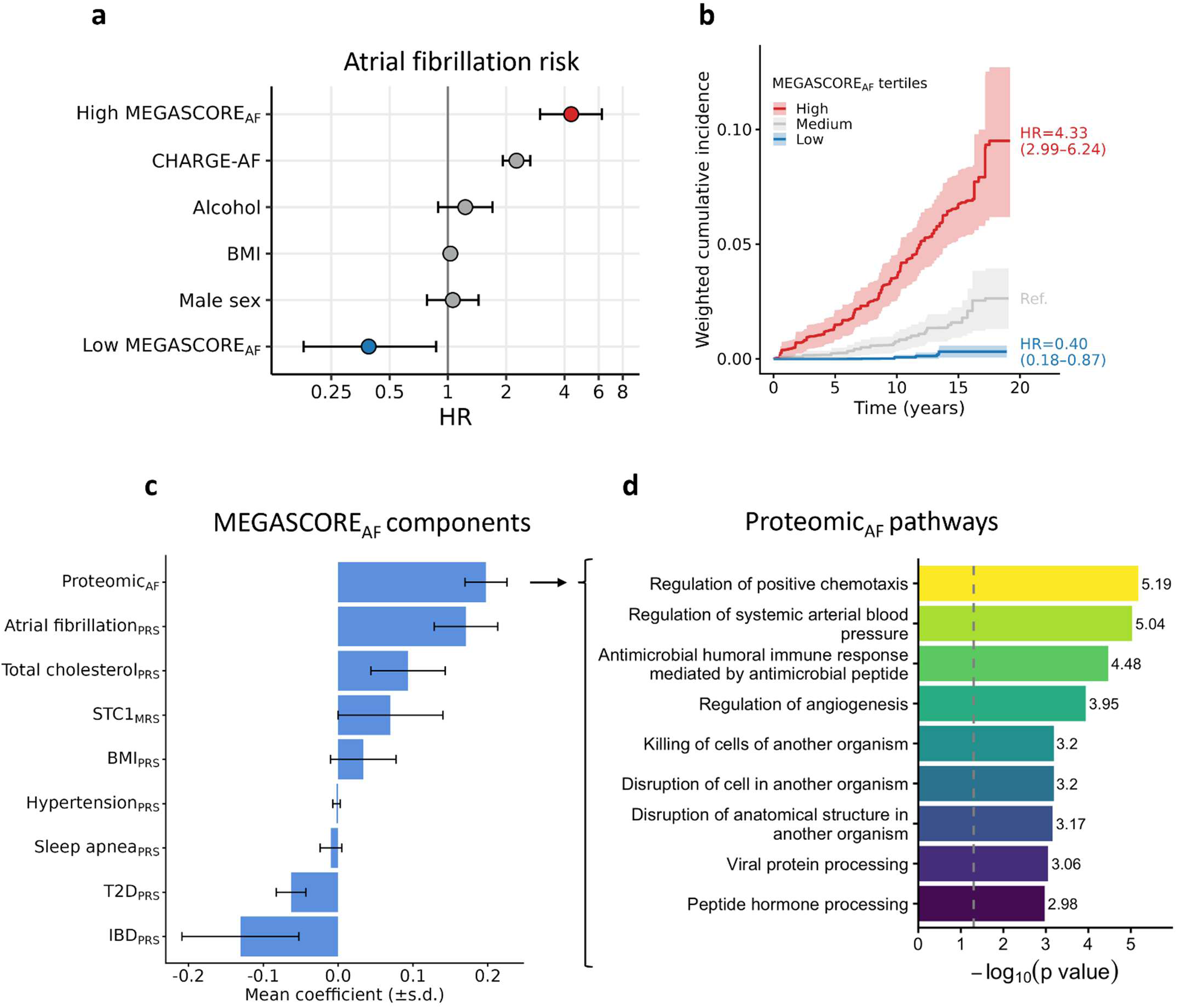
MEGASCOREs and biological atrial fibrillation risk. **a**, MEGASCORE_AF_ tertiles (high, medium [reference], and low) and future AF risk, adjusted for sex, BMI, alcohol consumption and the CHARGE-AF score. CHARGE-AF=Clinical risk score for incidence of atrial fibrillation composed of age, race, height, weight, systolic and diastolic blood pressure, smoking, antihypertensive therapy, diabetes, prior MI and HF. **b**, Cumulative incidence curves with 95% confidence intervals showing onset of AF over time when comparing individuals with high MEGASCORE_AF_, individuals with low MEGASCORE_AF_ and normal MEGASCORE_AF_. HRs and 95% confidence intervals from **a** are shown. **c**, Individual component scores contributing to the MEGASCORE_AF_, ordered by effect size. **d**, Gene Ontology enrichment analysis of proteins contributing to the proteomic component, highlighting significantly enriched biological pathways (-log_10_(P) values are shown). Abbreviations: PRS=polygenic score, MRS=methylation risk score, T2D=type 2 diabetes, STC1= stanniocalcin-1, IBD=inflammatory bowl disease, SD=standard deviation, HR=hazard ratio.

### MEGASCORE_AF_ and biological atrial fibrillation risk

High MEGASCORE_AF_ conferred a higher risk of AF (HR, 4.33; 95% CI, 3.00-6.24), and low MEGASCORE_AF_ values conferred a reduced risk of AF (HR, 0.39; 95% CI, 0.18-0.87), independent of clinical factors and the CHARGE-AF score (**Figure 5a,b**). The strongest contributors of the MEGASCORE_AF_ were the proteomic score for AF (Proteomic_AF_), the AF polygenic risk score (AF_PRS_), and total cholesterol polygenic risk score (**Figure 5c**). Pathway enrichment analyses of the proteins included in the Proteomic_AF_ score revealed significant overrepresentation of pathways related to hemodynamic stress, neurohormonal activation, vascular remodeling, and inflammatory signaling (**Figure 5c**). Corresponding downstream analyses for the other MEGASCOREs are provided in **Supplementary Figures 9-11**.

### Associations of MEGASCORE-derived phenotypic clusters with cardiovascular disease risk

Unsupervised MEGASCORE clustering identified four phenotypes with distinct cardiovascular risk profiles (**Figure 1d**). Participants in the global high-risk cluster exhibited the highest weighted incidence rates across all cardiovascular diseases, including AF (0.33 per 100 PY), MI (0.83 per 100 PY), HF (0.32 per 100 PY), and cardiovascular mortality (1.24 per 100 PY) (**Figure 6a**). In contrast, the low-risk cluster consistently showed low incidence across all diseases (0.02-0.09 per 100 PY). In Cox analyses for the composite cardiovascular disease outcome, the risk was highest in the global high-risk cluster (HR, 4.91; 95% CI, 3.57-6.75; P=1.01×10^−22^), followed by the mixed cardiovascular (HR, 2.06; 95% CI, 1.55-2.74; P=6.20×10^−7^) and AF-enriched cluster (HR, 1.87; 95% CI, 1.44-2.44; P=6.20×10^−7^; **Figure 6b**).

**Figure 6.**
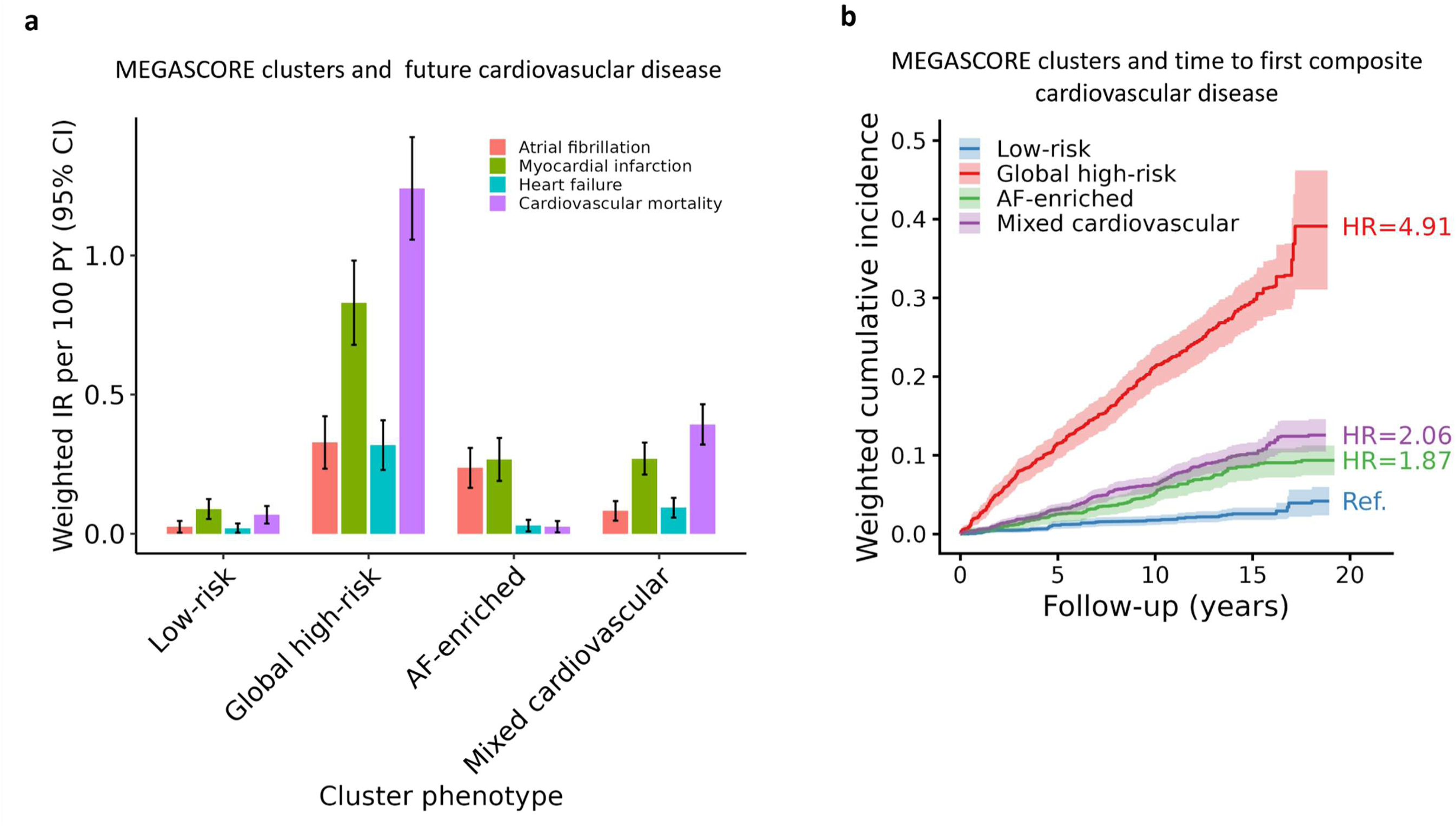
MEGASCORE phenotype clusters and cardiovascular disease risk. **a**, Incidence rates per 100 person-years for cardiovascular diseases across four MEGASCORE-derived phenotype clusters (low-risk, global high-risk, AF-enriched, mixed cardiovascular risk). **b**, Cumulative incidence curves for time to first occurrence of the composite cardiovascular disease endpoint (AF, HF, MI, or cardiovascular mortality) across clusters. Hazard ratios were estimated using multivariable Cox regression adjusted for age, sex, ancestry, BMI, smoking, height, weight, hypertension, diabetes, and hypercholesterolemia, with the low-risk cluster as the reference group.

## Discussion

In this study, we developed 4 cardiovascular disease-specific multiomic scores integrating proteomic signatures, polygenic scores, and DNA methylation scores to characterize risk of MI, AF, HF, and cardiovascular mortality. The MEGASCOREs were associated with their respective outcomes after adjustment for conventional cardiovascular risk factors. They also provided prognostic information among participants without conventional risk factors and demonstrated consistent risk stratification across ancestry groups. Together, these findings support the potential value of integrating multiple molecular layers to characterize cardiovascular risk beyond conventional clinical assessment.

The relative contributions of the 3 omic layers differed across outcomes. Polygenic scores accounted for a larger proportion of components in the MEGASCORE_MI_ and MEGASCORE_AF_, both of which have substantial inherited components.^30–33^ Proteomic scores contributed more to the MEGASCORE_HF_ and DNA methylation scores to the MEGASCORE_CV-Death_. Together, these results underscore that the molecular architecture of cardiovascular disease risk spans both static (genetic) and dynamic (proteomic and epigenetic) biological layers.

An important finding was that MEGASCOREs remained associated with cardiovascular disease outcomes among individuals without conventional risk factors. This observation suggests that molecular profiles may capture risk not identified by routine clinical assessment. Such information could complement established prevention strategies, particularly in individuals whose clinical risk profiles suggests low risk.^34^ However, the presence of a prognostic association does not establish that molecular screening improves clinical outcomes or that preventive treatment should be initiated based on these scores. Their clinical value will depend on reproducibility, actionable thresholds, and evidence that scores-guided management improves outcomes.

We also evaluated the MEGASCORE across ancestry groups. Discrimination for cardiovascular mortality was higher for MEGASCORE_CV-Death_ than for SCORE2 in ancestry-specific comparisons, and calibration analyses suggested differences in performance across ancestries for the scores. Risk prediction models developed primarily in European populations often require recalibration when applied to other populations because of differences in risk factor distributions and disease incidence.^27,35,36^ The robust cross-ancestral performance of MEGASCORE_CV-Death_ indicates that integrated multiomic profiles may capture biological processes that are more broadly shared across populations.

The lifestyle analyses provide additional context for interpreting the MEGASCOREs. Favorable lifestyle profiles were associated with lower MEGASCORE values, particularly for MEGASCORE_MI_ and MEGASCORE_CV-Death_. This suggests that some of the molecular variation reflected by scores relates to modifiable exposures. This observation raises the possibility that favorable lifestyle behaviors may attenuate molecular risk,^20,37^ although whether MEGASCOREs are modifiable over time warrants further investigation.

The MEGASCOREs allowed us to characterize participants according to their combined molecular risk profiles. The global high-risk cluster had the highest observed incidence rates across cardiovascular disease outcomes, whereas the low-risk cluster showed the lowest rates. The AF-enriched and mixed cardiovascular clusters showed intermediate patterns. Previous studies have demonstrated that molecular clustering can identify prognostically distinct subgroups that are not distinguished by conventional clinical variables.^38–40^ Our findings extend this concept by integrating disease-specific scores across a broad spectrum of cardiovascular disease. Further work is needed to determine whether these molecular cluster profiles inform clinical decision-making.

The MEGASCOREs provide insights into the biological mechanisms underlying cardiovascular disease. For example, the MEGASCORE_AF_ prominently featured the methylation score for stanniocalcin-1 (SCT1), a protein implicated in cardiac fibrosis and calcium transport, processes potentially relevant to arrhythmogenesis.^41,42^ Other contributors included polygenic risk score for BMI and total cholesterol. Consistent with previous observational and genetic studies, obesity is an established causal risk factor for AF,^43,44^ whereas higher total cholesterol levels have paradoxically been associated with lower AF risk.^45^ These findings illustrate how integrative multiomics can capture biologically relevant pathways associated with cardiovascular disease.

Our study has some potential limitations. First, the ancestry-specific analyses may have limited precision in smaller subgroups, and additional validation is needed in populations not adequately represented in the cohorts. Second, the predictive performance of the MEGASCOREs does not establish clinical utility. Third, the observational associations cannot establish that changes in the scores mediate reductions in cardiovascular outcomes. Finally, the associations between MEGASCOREs and cardiovascular disease outcomes are observational and should not be interpreted as causal without additional mechanistic or genetic evidence.

In summary, the MEGASCORE framework integrates genomic, epigenomic, and proteomic information to characterize risk across multiple cardiovascular diseases. The scores provided prognostic information beyond conventional clinical variables, including among individuals without risk factors. Prospective evaluation of clinical utility is needed to determine whether integrated multiomic profiling can improve cardiovascular prevention.

## Data availability statement

UK Biobank data are available upon request to qualified researchers through a standard protocol (https://www.ukbiobank.ac.uk/register-apply). Individual-level data from PURE, G-CHF or ORIGIN are not available to the public because participants did not consent to the sharing of their personal data outside the investigative team. Formal proposals for collaboration with other groups with similar data for pooled analyses will be considered.

## Code availability

All statistical analyses and figure generation were performed in R (v4.4.2) using publicly available packages, including glmnet, pROC, caret, ggplot2, survival, epitools, purrr, DescTools, clusterProfiler, and org.Hs.eg.db. Custom code used for MEGASCORE derivation is available in the megascore GitHub repository (https://github.com/Meyrep/megascore).

## Acknowledgements

The PURE study is an investigator-initiated study that is funded by the Population Health Research Institute, the Canadian Institutes of Health Research (CIHR), Heart and Stroke Foundation of Ontario, support from CIHR’s Strategy for Patient Oriented Research, through the Ontario SPOR Support Unit, as well as the Ontario Ministry of Health and Long-Term Care and through unrestricted grants from several pharmaceutical companies (with major contributions from AstraZeneca [Canada], Sanofi-Aventis [France and Canada], Boehringer Ingelheim [Germany and Canada], Servier, and GlaxoSmithKline), and additional contributions from Novartis and King Pharma and from various national or local organizations in participating countries as follows: Argentina—Fundacion ECLA; Bangladesh-Independent University, Bangladesh and Mitra and Associates; Brazil—International Research Center, Hospital Alemão Oswaldo Cruz, São Paulo, Brazil; Canada—Public Health Agency of Canada and Champlain Cardiovascular Disease Prevention Network; Chile—Universidad de la Frontera; Colombia—Colciencias (grant number 6566–04–18062); South Africa—The North-West University, SANPAD (SA and Netherlands Programme for Alternative Development), National Research Foundation, Medical Research Council of South Africa, The South Africa Sugar Association, Faculty of Community and Health Sciences; Sweden—grants from the Swedish State under the Agreement concerning research and education of doctors, the Swedish Heart and Lung Foundation, the Swedish Research Council, the Swedish Council for Health, Working Life and Welfare, King Gustaf V’s and Queen Victoria Freemasons Foundation, AFA Insurance, Swedish Council for Working Life and Social Research, Swedish Research Council for Environment, Agricultural Sciences and Spatial Planning, grant from the Swedish State under (LäkarUtbildningsAvtalet) Agreement, and grant from the Västra Götaland Region; and United Arab Emirates—Sheikh Hamdan Bin Rashid Al Maktoum Award for Medical Sciences, Dubai Health Authority, Dubai. The PURE biomarker project was supported by Bayer and the CIHR. The biomarker project was led by PURE investigators at the Population Health Research Institute (Hamilton, Canada) in collaboration with Bayer scientists. Bayer directly compensated the Population Health Research Institute for measurement of the biomarker panels, scientific, methodological, and statistical work, but had no role in the interpretation of the results or development of the manuscript. Genetic analyses were supported by CIHR (G-18–0022359) and Heart and Stroke Foundation of Canada (application number 399497) in the form of funding to GP. The G-CHF study was supported by funding from Bayer and the Canadian Institutes of Health Research (CIHR). The ORIGIN trial and biomarker project were supported by Sanofi and CIHR. The biomarker project was led by ORIGIN investigators at the Population Health Research Institute (Hamilton, Canada) with the active collaboration of Sanofi scientists. Sanofi directly compensated Myriad RBM, Inc. for measurement of the biomarker panel and the Population Health Research Institute for scientific, methodological and statistical work. This project was also supported by the Innovative Medicines Initiative 2 Joint Undertaking under grant agreement no. 115974 (BEAt-DKD). This Joint Undertaking receives support from the European Union’s Horizon 2020 Research and Innovation Programme, the European Federation of Pharmaceutical Industries and Associations, and JDRF. Genetic analysis of ORIGIN participants was supported by CIHR (award 125794 to G.P.).

